# Industry ties and evidence in public comments on the FDA framework for modifications to artificial intelligence/machine learning-based medical devices: a cross sectional study

**DOI:** 10.1101/2019.12.11.19013953

**Authors:** James A Smith, Roxanna E Abhari, Zain U Hussain, Carl Heneghan, Gary S Collins, Andrew J Carr

**Affiliations:** Botnar Research Centre, Nuffield Department of Orthopaedics, Rheumatology and Musculoskeletal Sciences, University of Oxford, UK; National Institute for Health Research Oxford Biomedical Research Centre, John Radcliffe Hospital, Oxford, UK; College of Medicine and Veterinary Sciences, University of Edinburgh, UK; Centre for Evidence-Based Medicine, Nuffield Department of Primary Care Health Sciences, University of Oxford, UK; Centre for Statistics in Medicine, Nuffield Department of Orthopaedics, Rheumatology and Musculoskeletal Sciences, University of Oxford, UK

**Keywords:** conflict of interest, financial interest, policy, regulation, financial ties

## Abstract

**Objectives:** To determine the extent and disclosure of financial ties to industry and use of scientific evidence in comments on a US Food and Drug Administration (FDA) regulatory framework for modifications to artificial intelligence/machine Learning (AI/ML)-based software as a medical device (SaMD).

**Design:** Cross-sectional study.

**Setting:** We searched all publicly available comments on the FDA “Proposed Regulatory Framework for Modifications to Artificial Intelligence/Machine Learning (AI/ML)-Based Software as a Medical Device (SaMD) - Discussion Paper and Request for Feedback” from April 2^nd^ 2019 to August 8^th^ 2019.

**Main outcome measures:** The proportion of articles submitted by parties with financial ties to industry, disclosing those ties, citing scientific articles, citing systematic reviews and meta-analyses, and using a systematic process to identify relevant literature.

**Results:** We analysed 125 comments submitted on the proposed framework. 79 (63%) comments came from parties with financial ties; for 36 (29%) comments it was not clear and the absence of financial ties could only be confirmed for 10 (8%) comments. No financial ties were disclosed in any of the comments that were not from industry submitters. The vast majority of submitted comments (86%) did not cite any scientific literature, just 4% cited a systematic review or meta-analysis, and no comments indicated that a systematic process was used to identify relevant literature.

**Conclusions:** Financial ties to industry were common and undisclosed and scientific evidence, including systematic reviews and meta-analyses, were rarely cited. To ensure regulatory frameworks best serve patient interests, the FDA should mandate disclosure of potential conflicts of interest (including financial ties), in comments, encourage the use of scientific evidence and encourage engagement from non-conflicted parties.

**Strengths and limitations of this study:**

- We analysed the extent of financial ties to industry and the use of scientific evidence in comments on the proposed FDA framework
- We used a comprehensive strategy to attempt to identify financial ties to industry
- Readers may be able to contribute higher quality comments to subsequent drafts of this framework
- There is heterogeneity in the degree of conflict with respect to the framework that the recorded financial ties may represent; some ties will be more likely than others to result in biased commenting
- Because the framework could not be classified as pro-industry or not, we did not classify the direction of opinions expressed in comments with respect to the framework and their association with financial ties
- We do not know how information submitted to FDA is used internally in the rule-making process

## Introduction

Artificial intelligence and machine learning (AI/ML) are increasingly prevalent in the healthcare literature^1^. At least 14 medical devices incorporating AI/ML have now been cleared by the US FDA^2^, including an autonomous diagnostic system for diabetic retinopathy that does not require input from a clinician for interpretation^3^. Because artificial intelligence may learn and adapt to additional data in real time to improve performance, regulators have questioned the suitability of traditional medical device regulatory pathways for AI/ML containing devices. Changes that might affect a device’s performance under the current regulatory framework would require further review from the FDA, which is time-consuming and may not suit the iterative modification that often characterises software development and deployment. The FDA has therefore proposed a regulatory framework for modifications to AI/ML-based SaMD^4^ (Box 1). Because the FDA is one of the most prominent regulatory agencies, other agencies may follow FDA regulatory approaches. It is therefore essential that the framework reflects and promotes patient interests and safety.

### Box 1

*Summary of AI/ML framework: “Proposed Regulatory Framework for Modifications to Artificial Intelligence/Machine Learning (AI/ML)-Based Software as a Medical Device (SaMD) - Discussion Paper and Request for Feedback”*^4^

The FDA released the first discussion paper on April 2^nd^ 2019 outlining a framework for regulating modifications to SaMD that use Artificial Intelligence (AI) and Machine Learning (ML). The comment period closed on June 3^rd^ 2019. The proposed framework describes:

- The extent to which FDA’s traditional framework for assessing modifications to devices should apply to AI/ML SaMD
- Modification categories for continuously learning AI/ML SaMD and a proposed “pre-determined change control plan” in pre-market submissions: when seeking regulatory approval, manufacturers would also submit a plan for modifications, including model retraining, as part of the initial pre-market review.
- Expectations for manufacturers to monitor the real-world performance of AI/ML systems and periodically report updates to users and to the FDA on what changes have been implemented.
- The evaluation, monitoring, and management of risks from AI/ML modifications from initial pre-market submission through to post-market performance.
- Hypothetical examples of modifications and their applicability to the proposed framework.

Agencies base decisions on sound reasoning and scientific evidence^5^, and the process of developing FDA regulations and guidance involves opportunities for the public to assess and comment on proposed rules before they are finalised. Comments can be submitted by anyone and are considered by the FDA in subsequent drafts and final rulings. However, there is potential for financial conflicts of interest (COI) among commenters, who could serve to benefit from particular outcomes such as less stringent regulatory requirements. Therefore, we evaluated the prevalence and disclosure of financial ties to industry in comments on the recent proposed AI/ML device framework. There is a huge academic literature available related to AI/ML, which could be used to inform the development of new regulations, and the FDA explicitly look for ‘good science’ in submitted comments^6^. We also, therefore, examined the citation of scientific evidence, including of systematic reviews, to determine whether there is opportunity to increase and improve its use in comments.

## Methods

The docket folder for comment submission for the “Proposed Regulatory Framework for Modifications to Artificial Intelligence/Machine Learning (AI/ML)-Based Software as a Medical Device (SaMD) - Discussion Paper and Request for Feedback” ^7^ was accessed 8^th^ August 2019 and meta-data for all comments exported to Microsoft Excel. Any comments submitted after this date, which was after the comment period closed, were not included in analysis. Individual comments were subsequently accessed via the docket by following the link in the Excel export. We conducted a pilot study in which JAS developed a data extraction protocol by reading and preliminarily analysing all comments submitted on the proposed framework. Two reviewers (REA and ZUH) then independently extracted data from all comments according to the extraction protocol (available on Open Science Framework: https://osf.io/g423d/). We accessed the data from August to October 2019 and the comment order was randomised for each reviewer. JAS consolidated any discrepancies between the reviewers. Some minor changes were made to the data extraction procedure during the consolidation process which are described in the extraction protocol. JAS extracted and checked any new data as required.

### Category

Comment submitters were categorised according to the categories in Table 1. Some submitters provide a ‘Category’ in the ‘Submitter Information’ section of the docket folder. When the category could not be determined by the extractor, the category provided in the submitter information was used, if provided.

**TABLE 1:** CATEGORIES OF SUBMITTERS AND EXPLANATIONS

| Category | Explanation |
| --- | --- |
| *Academia* | Included individual academics and academic groups |
| *Healthcare* | Included healthcare associations and health professionals |
| *Industry* | Included companies, industry associations and individuals from industry |
| *Individual consumers* | Category only used when provided in the submitter information |
| *Mixed associations* | Defined as associations comprising at least one industry and non-industry organisation |
| *Federal government* | Category only used when provided in the submitter information |
| *Spam* | Recorded when the comment content was clearly not related to the content of the FDA document |
| *Other* | Recorded when the submitter was not one of the above categories but was identifiable. An example was the US Technology Policy Committee of the Association for Computing Machinery |
| *Unknown* | Recorded when no category was listed in the submitter information, insufficient information was provided to identify the submitter category (i.e. a name, but no affiliation was provided) and when the commenter was listed as “Anonymous”. |

### Financial ties to industry

We searched for financial ties among comment submitters. A financial tie was defined as a financial link with industry. Submitters in the industry or mixed association category were assumed to have a financial interest, For other submitters, we determined whether there were financial ties according to the following method:

1. If the comment submitter is was an academic (or group thereof), we searched the name of the academic(s) plus their institution to find academic pages or pages mentioning them on industry websites and determined if they were advisors or board members for industry, or had another obvious link. If this was not the case, we examined the two most recent publications (published from July 2017 until October 2020) that we could identify for each author and checked for industry affiliations and disclosures of personal fees, speaking fees, board-membership, employment, grants, or similar from industry or industry associations. If at least one author of the comment had a financial tie according to these criteria, we recorded that there was a financial tie. If, of the two most recent publications, at least one stated that there was no financial tie or conflict of interest to disclose (or similar wording), and the other did not disclose a financial tie, we recorded no financial tie. If only one paper was identifiable, this was deemed sufficient to identify a conflict or lack thereof (i.e. if only one paper was found and this stated there was no conflict, we recorded ‘no’).
2. For individual consumers or health professionals that provided their institution or another means of identifying them, we followed step 1 and additionally searched the open payments database (https://openpaymentsdata.cms.gov/) for their name and looked for contributions from industry of any sort form the last two years. A financial tie was recorded if the submitter had received contributions from industry and we were able to verify that the individual in the database was the commenter, for example by cross-referencing their institution.
3. For healthcare associations and ‘other’ submitters, we searched for financial ties according to step 2 among all of the authors listed on the comment and, if no authors were listed, among board members. If any authors, or at least half of the board members had financial ties, the submitter was considered to have a financial tie. When no financial tie could be identified for an association, we recorded that there was none.
4. For submitters in the federal government category, we assumed that there was no financial tie.
5. When the presence or absence of a financial tie could not be determined according to these criteria, it was recorded as unknown.

### Disclosure of financial ties

For industry submitters, we assumed that disclosure would not be required because the potential COI is self-evident, but that for any other potentially conflicted submitters a disclosure would be required for the FDA to be aware of the financial tie. Therefore, for non-industry submissions with financial ties, we recorded whether or not ties were disclosed.

### Scientific evidence and systematic searches

As a proxy for the use of scientific evidence in comments, we recorded whether comments cited scientific articles, systematic reviews or meta-analyses, and whether any systematic search was reported to identify any documents or literature. We identified articles by reading each comment and examining footnotes, bibliographies or in-text citations, and considered academic journal articles or pre-prints (for example, papers on ArXiv or IEEE) to be scientific articles. To identify citations of systematic reviews or meta-analyses, we looked at the titles of any referenced articles for the terms (or similar terms to) “systematic review” or “meta-analysis” and, where it was unclear, we reviewed the abstract or if necessary the full-text. We determined whether or not a systematic search was used to identified documents or literature referenced or discussed in the comment. A systematic search in this context refers to a process that would be repeatable by an independent reader, such as listing the search terms used to identify literature.

### Document length

We recorded the length of each comment to give an indication of its comprehensiveness. Specifically, we recorded the length, in pages, of any of the attachments on the docket page for the comment. If multiple attachments were provided, we recorded sum of the lengths, (unless attachments were duplicates, for example, one in word and one as a pdf, in which case we recorded non-duplicates). For comments that did not provide an attachment, we copied and pasted the comment into a word document with standard font size 12 and recorded the length in pages.

### Data analysis

Duplicate, near duplicate and spam comments were excluded from analysis. All other comments were included. Analysis was descriptive and was conducted in R v3.6.1 [ref: ^8^]. The R code and dataset are provided on Open Science Framework (https://osf.io/g423d/). The analysis was reproduced in Python independently by ZUH.

### Patient and public involvement

There was no patient and public involvement in this study.

## Results

130 public comments were submitted in response to the proposed regulatory framework at the time we downloaded data (8^th^ August 2019), of which 5 were duplicates, near-duplicates, or spam, leaving 125 comments for analysis. Combining industry, mixed associations (which include representatives of industry) and other contributors with financial ties revealed that at least 79 (63%) comments came from parties with financial ties (Table 2). For 36 comments, (29%) it was not clear, and the absence of a financial ties could be confirmed in only 10 comments (8%). The length of comments and proportion of comments citing scientific evidence was similar across submitters regardless of financial interest (Table 2).

**TABLE 2:** PRESENCE OF FINANCIAL INTERESTS, CITATION OF SCIENTIFIC EVIDENCE, AND DOCUMENT LENGTH

| **Financial ties to industry** | **% of total (n)** | **% citing scientific evidence (n)<sup>1</sup>** | **Median document length, pages (range)** |
| --- | --- | --- | --- |
| Yes<sup>2</sup> | 63.2% (79) | 16.5% (13) | 4 (1-30) |
| No | 8% (10) | 10% (1) | 2.5 (1-13) |
| Unknown | 28.8% (36) | 8.3% (3) | 1 (1-7) |

<sup>1</sup>Including articles in journals or preprints
<sup>2</sup>Includes industry, industry associations, mixed associations, and any other submitters for which we were able to identify a financial tie to industry

Table 3 summarises the categories of organisations or individuals submitting comments. Industry submitted 64 (51%) comments. Of the 61 non-industry comments, we were able to determine whether or not there were financial ties to industry in 25 submissions (41%). Of these 25, financial ties for 15 were identified, and none were found for 10. No financial ties were disclosed in any of the non-industry comments.

**TABLE 3:** CATEGORY OF ORGANISATION OR INDIVIDUAL SUBMITTING COMMENT, CITATION OF SCIENTIFIC EVIDENCE, AND DOCUMENT LENGTH

| **Category<sup>1</sup>** | **% of total (n)** | **% citing scientific evidence (n)<sup>2</sup>** | **Median document length, pages (range)** |
| --- | --- | --- | --- |
| Industry<sup>2</sup> | 51.2% (64) | 12.5% (8) | 4 (1-30) |
| Academia<sup>3</sup> | 17.6% (22) | 9.1% (2) | 1 (1-13) |
| Healthcare<sup>4</sup> | 9.6% (12) | 16.7% (2) | 2.5 (1-8) |
| Unknown | 7.2% (9) | 0% (0) | 1 (1-6) |
| Other | 5.6% (7) | 57.1% (4) | 2 (1-5) |
| Mixed association<sup>5</sup> | 4% (5) | 20% (1) | 5 (4-8) |
| Individual consumer | 3.2% (4) | 0% (0) | 1 (1) |
| Federal government | 1.6% (2) | 0% (0) | 1.5 (1-2) |

<sup>1</sup>See Table 1 for explanations of categories
<sup>2</sup>Percentage of that “Category” citing scientific evidence, including articles in journals or preprints

Of the 125 comments, 108 (86%) did not cite scientific literature, 15 (12%) cited at least one paper published in an academic journal, and two (1.6%) cited article pre-prints but not papers in academic journals. Five (4%) comments cited a systematic review or meta-analysis, and none of the 125 comments reported any systematic method for identifying the literature referenced in the document.

## Discussion

### Statement of principal findings

Industry and other submitters with financial ties to industry comprised nearly two thirds of the comments on the proposed FDA framework. We found no evidence that submitters without a financial interest were more likely to cite scientific evidence or contribute more comprehensive comments. Identifying financial ties was not straightforward and required extensive searching. For many comments, the presence of financial ties was unknown: it is not possible to identify financial ties for anonymous commenters, or those that provide a name but no organisation, for example. The absence of financial ties could be confirmed in very few cases and disclosure of ties was non-existent. Scientific literature was rarely cited across all submissions, just five comments cited systematic reviews and meta-analyses, and no comments reported a systematic process for identifying the literature. There were far fewer academic submissions than those from industry.

### Strengths and limitations

In terms of strengths, this study is the first, that we know of, to examine financial ties among commenters on a proposed FDA regulatory framework, and adds to the growing literature documenting the prevalence of industry interests in public comment processes^9^. We used a thorough method for identification of financial ties which has identified more potentially conflicted parties than would be apparent from the self-identification of the commenters. We also examined the citation of scientific evidence, adding to literature documenting lack of scientific evidence in public comments in other contexts^10^. The raw data from this study is available publicly, so that readers can view specific financial ties, evidence cited, or reanalyse the data if desired. Finally, the AI/ML regulatory framework is still under development, and these findings may motivate better disclosure and use of scientific evidence as it continues to be developed.

In terms of limitations, the prevalence of financial ties in our study (63%) is likely to be an underestimate. For 29% (36) of commenters we were unable to confirm the presence or absence of financial ties because insufficient information was provided to identify their presence or absence according to our search strategy. Therefore, 63% should be interpreted as an estimate of the minimum prevalence of industry ties. In addition, there is heterogeneity in the degree of conflict with respect to the framework that the recorded financial ties may represent; some ties will be more likely than others to result in biased commenting.

Relatedly, we did not classify the direction of opinions expressed in comments with respect to the framework and their association with financial ties, in contrast to other work in similar areas^11,12^. Because of the early stage and diversity of topics proposed in the regulatory framework, it is not clear that supporting the framework would be putting financial considerations ahead of patient considerations, or vice versa. We are, therefore, only able to make any inferences regarding the prevalence of financial ties, and not their impact on the opinions expressed in comments with regards to considerations of health vs. financial interests. This is an important limitation because we cannot assume that financial ties will lead to biased commenting or will necessarily represent COI. In some cases, the presence of industry ties or COI do not introduce bias and may be aligned with patient interest^13,14^; however, in others, there is ample evidence that they do lead to bias^11,12,15–17^.

We have access only to the information externally submitted to FDA by the public, and undoubtedly further evidence is considered internally. We also do not know how the information gathered from public consultations is used internally and how it will impact the future regulatory framework. Agencies do not make decisions simply based on a majority of votes^5,9^, so prevalence of financial ties and lack of use of scientific evidence does not necessarily mean that decision making internally is biased. However, the purpose of public consultations is to gather information from the public to inform regulation. It is important that such information is high quality, reasoned, objective and transparent, where possible.

### Implications and recommendations

We recommend that the FDA requests a statement of interests in comments, which would help to further determine the extent of COI in future public consultations. They could also reject anonymous comments unless there is a clear reason that anonymity would improve comment quality. Currently, the regulations.gov ‘Tips for submitting effective comments’ document does not mention disclosure of COI or financial ties^5^, nor does the FDA’s information page on submitting comments^6^. In the absence of this requirement, submitters should proactively disclose any conflicts of interest. COI may influence drug approvals^15^, policy making^11,12^ and reporting of results in research^16,17^, and may therefore also be influential in the present context. Although disclosure will not necessarily change the interpretation of information, it is an important first step in improving transparency.

We encourage greater participation by non-conflicted parties and academics in the development of this framework in the future. Participation should come both from experts in the technology, as well as from the broader medical community that will use and evaluate it. FDA could proactively engage with, for example, journals with expertise in the relevant areas to notify readers when relevant legislation is drafted. However, the responsibility should not lie solely with FDA, but participation in the commenting process can also be driven and rewarded by academic departments or groups with relevant expertise. Continued input from associations and the greater use of scientific evidence, in particular that from systematic reviews, by all commenters, could be valuable. At present, there are few relevant systematic reviews for this purpose (though exceptions include ref: ^18^) and further work in this area would be useful.

In other areas of medical device regulation, there is literature available evaluating or critiquing regulatory frameworks^19–21^. However, once regulations have been developed it is a laborious process for them to be changed. AI/ML regulation is in an early stage where scientific knowledge could be used prospectively to help to define an appropriate framework and reduce the probability of issues occurring in the future. The regulations.gov guidance on commenting states: “Although public support or opposition may help guide important public policies, agencies make determinations for a proposed action based on sound reasoning and scientific evidence rather than a majority of votes. A single, well-supported comment may carry more weight than a thousand form letters”^5^. If high quality comments are provided in future developments of the proposed rules, supported by sound scientific evidence, they may well be weighted more highly in decision making than other comments and directly impact the framework under development.

### Conclusion

We found that the prevalence of financial ties to industry in commenters was high. For nearly 30% of comments, we were unable to determine whether or not there was a financial tie, and disclosure of ties was non-existent. The proportion of academic submitters was relatively low, and the use of scientific evidence to support comments was sparse. We recommend that the FDA require disclosure of potential COI, and encourage greater academic participation and use of scientific evidence in public comments. The generalizability of this work has not been determined, and future work could extend the investigation to other policy areas or jurisdictions.

## Data Availability

The data, code, and data collection protocol are available on open science framework: https://osf.io/g423d/

## Author contributions

JAS conceived the study, conducted a pilot study, analysed the data in R and wrote the first draft of the manuscript and is the guarantor. REA extracted data from public comments and contributed to the first draft. ZUH extracted data from public comments and repeated the data analysis independently in Python. All authors interpreted the data, reviewed and critically appraised the manuscript for important intellectual content. The corresponding author attests that all listed authors meet authorship criteria and that no others meeting the criteria have been omitted.”

## Data sharing

Data, code and the data extraction protocol are publicly available at https://osf.io/g423d/.

## Ethics approval

Ethics approval was not required.

## Transparency statement

The lead author (JAS) affirms that the manuscript is an honest, accurate, and transparent account of the study being reported; that no important aspects of the study have been omitted; and that any discrepancies from the study as originally planned (and, if relevant, registered) have been explained.

## Patient and public involvement statement

There was no patient and public involvement in this study.

## Dissemination declaration

Dissemination to study participants or patient organisations is not applicable.

